# Temporal Patterns of Antithrombotic Therapy and Clinical Outcomes After Atrial Fibrillation-Related Stroke

**DOI:** 10.1101/2025.08.01.25332685

**Authors:** Keon-Joo Lee, Hoonji Oh, Han-Gil Jeong, Beom Joon Kim, Moon-Ku Han, Joon-Tae Kim, Kang-Ho Choi, Dong-Ick Shin, Jae-Kwan Cha, Dae-Hyun Kim, Dong-Eog Kim, Jong-Moo Park, Kyusik Kang, Soo Joo Lee, Jae Guk Kim, Mi-Sun Oh, Kyung-Ho Yu, Byung-Chul Lee, Keun-Sik Hong, Yong-Jin Cho, Jay Chol Choi, Tai Hwan Park, Kyungbok Lee, Jee-Hyun Kwon, Wook-Joo Kim, Jun Lee, Ji Sung Lee, Da Young Hong, Juneyoung Lee, Philip B. Gorelick, Hee-Joon Bae, CRCS-K (Clinical Research Collaboration for Stroke in Korea) investigators

## Abstract

**Background:** There is clinical equipoise in relation to the optimal timing and strategy for antithrombotic therapy in acute ischemic stroke (AIS) patients with atrial fibrillation (AF) who may be eligible for non-vitamin K antagonist oral anticoagulants (NOACs). This study aimed to describe the temporal evolution of antithrombotic treatment strategies and associated clinical outcomes after AF-related AIS.

**Methods:** This multicenter prospective cohort study enrolled 2,965 patients with AIS and AF from 16 tertiary stroke centers across South Korea between February 2018 and January 2021, with follow-up completed by January 2022. The primary outcome was a composite of recurrent stroke, myocardial infarction, and all-cause death. Secondary outcomes included individual components of the primary outcome and major bleeding events.

**Results:** The median follow-up duration was 1.92 years. Among 2,965 patients (mean [SD] age, 75.3 [10.2] years; 54.1% male), antithrombotic strategies varied widely in the acute phase. Within 48 hours of admission, 50.9% received antiplatelet-only therapy (29.4% single, 21.5% dual), 23.2% received NOAC monotherapy, and 10.4% received a combination of antiplatelets and anticoagulants. By discharge, NOAC monotherapy had become the predominant treatment strategy (65.7%), and this pattern persisted throughout follow-up.

This treatment heterogeneity coincided with distinct temporal patterns in clinical outcomes. The incidence of the primary endpoint was highest in the first two weeks (32.70 [95% CI, 29.64-36.06] per 100 person-months), and declined thereafter. Across all time periods, patients receiving NOAC monotherapy consistently had lower incidence rates (3-month rate: 4.95 [95% CI, 4.37-5.61] per 100 person-months) than those receiving antiplatelet-only therapy (11.98 [95% CI, 9.57-15.01]) or no antithrombotic therapy (18.44 [95% CI, 14.26-23.86]).

**Conclusions:** In this prospective cohort of patients with AF-related stroke, early antithrombotic treatment strategies were heterogeneous but evolved primarily toward use of NOAC monotherapy by the time of discharge from the hospital and during longitudinal follow-up. Treatment selection was associated with marked differences in outcomes, particularly during the early high-risk period. These findings underscore the need for safe and effective, time-sensitive antithrombotic strategies, especially in patients for whom immediate anticoagulation is deferred.

**Clinical Perspective:** *What Is New?:*

- This nationwide prospective cohort study evaluated temporal trends in antithrombotic therapy and outcomes in patients with atrial fibrillation (AF)-related ischemic stroke.
- There was substantial heterogeneity in early treatment strategies, which converged toward NOAC (non–vitamin K antagonist oral anticoagulant) monotherapy by discharge.
- The risk of recurrent vascular events and death was highest during the first 2 weeks post-stroke and varied significantly by antithrombotic regimen.

*What Are the Clinical Implications?:*

- The early post-stroke period represents a critical window with elevated risk and variable treatment practices.
- NOAC monotherapy was consistently associated with the lowest event rates, supporting early initiation when feasible.
- Timely and individualized antithrombotic strategies are essential to improve outcomes, especially for patients in whom anticoagulation is initially deferred.

## Introduction

Non-vitamin K antagonist oral anticoagulants (NOACs) are a cornerstone of stroke prevention in patients with atrial fibrillation (AF).^1^ NOACs offer several advantages over vitamin K antagonists (VKAs) including predictable pharmacokinetics, rapid achievement of therapeutic drug levels, fewer drug-food interactions, lower rates of brain hemorrhage complications, and fixed dosing regimens that eliminate the need for routine coagulation monitoring.^2,3^ Their rapid onset and offset of action also confer practical benefits in the context of urgent surgical interventions and management of bleeding.^4,5^

Although NOACs have largely replaced warfarin in the management of AF-related stroke,^6–8^ several key questions remain unresolved. These include: 1) what antithrombotic strategies are implemented during the acute and chronic phases after stroke; 2) when are the risks of recurrent vascular events and bleeding complications highest; and 3) how do clinical outcomes vary by treatment strategy. While recent clinical trials have investigated the timing of NOAC initiation after acute stroke,^9,10^ the generalizability of these findings is limited by narrow inclusion criteria and short follow-up durations.

Such questions are difficult to address through randomized clinical trials alone, due to restrictions of patient eligibility criteria and duration of follow-up.^11,12^ Well-designed and implemented prospective registries can help to bridge this gap by capturing real-world treatment patterns across diverse populations.

To address these knowledge gaps, we conducted a nationwide, prospective cohort study of patients with AF-related acute ischemic stroke (AIS), with up to 3 years of follow-up. Our aim was to characterize contemporary antithrombotic treatment practices, examine how these strategies evolve over time after stroke onset, and evaluate temporal trends in key clinical outcomes in real-world settings.

## Methods

### Study Design and Population

This was a prospective cohort study of patients with atrial fibrillation (AF)-related AIS. Although the study population was derived from the Clinical Research Collaboration for Stroke in Korea (CRCS-K) registry,^13–15^ this investigation was conducted independently, with a separate protocol and informed consent process. Of the 20 CRCS-K participating centers, 16 tertiary stroke centers contributed to this cohort study. Patient enrollment occurred between February 2018 and January 2021.

Both the CRCS-K registry and this cohort study were approved by the institutional review boards of all participating centers, and written informed consent was obtained separately for each study.

Eligible patients were adults (age >18 years) who presented with AIS within 7 days from symptom onset and had either a prior diagnosis of AF or new-onset AF identified during hospitalization. All participants had diffusion-weighted imaging (DWI) evidence of acute ischemic lesions corresponding to their clinical symptoms. Patients who were unable to provide informed consent were excluded.

### Data Collection and Baseline Assessment

At enrollment, we collected demographic data and clinical characteristics, including stroke severity (National Institutes of Health Stroke Scale [NIHSS]), premorbid functional status (modified Rankin Scale [mRS]), and time from symptom onset to hospital arrival. Vascular risk factors and medical history—including hypertension, diabetes, hyperlipidemia, coronary heart disease, peripheral artery disease, current smoking status, and previous stroke or transient ischemic attack (TIA)—were also recorded. The presence of symptomatic steno-occlusion of relevant arteries was assessed at baseline using either CT or MR angiography.

Antithrombotic medication use was documented at predefined time points: prior to admission, within 48 hours of admission, at discharge, and during follow-up visits at 3 months, 1 year, 2 years, and 3 years. We recorded the specific types of agents used, including anticoagulants (NOACs, warfarin) and antiplatelet agents, as well as combination regimens. Medication adherence was assessed at the 1-year follow-up visit using self-reported compliance questionnaires.

### Outcome Measures and Follow-up

The primary outcome was a composite of stroke recurrence, myocardial infarction, and all-cause death. Secondary outcomes included individual components of the primary outcome, ischemic stroke recurrence, and bleeding events classified according to the Bleeding Academic Research Consortium (BARC) criteria, with major bleeding defined as type 3 or 5.^16,17^ Stroke recurrence included both new neurological deficits and progression of the index ischemic lesion within the same vascular territory, consistent with definitions used in prior clinical trials of minor ischemic stroke.^18–20^

Follow-up assessments were conducted at predefined intervals via in-person clinic visits or structured telephone interviews using standardized questionnaires. The final follow-up exams were completed in January 2022, ensuring a minimum follow-up of 1 year and a maximum of 3 years for all participants. All suspected outcome events were adjudicated by an independent clinical event committee blinded to patients’ antithrombotic treatment status.

### Statistical Analysis

Baseline characteristics were summarized using means and standard deviations or medians with interquartile ranges for continuous variables, and frequencies with percentages for categorical variables. Follow-up time was calculated from the date of symptom onset (designated as time zero).

Antithrombotic treatment was assessed at predefined intervals: within 48 hours of admission, at discharge, and at 3 months, 1 year, 2 years, and 3 years. Treatment regimens were initially categorized into 7 groups: no treatment, NOAC monotherapy, VKA monotherapy, other anticoagulants, single antiplatelet, dual antiplatelet, and combination of antiplatelet and anticoagulant agents. A Sankey diagram illustrated the transitions in treatment over time.

Group comparisons of patient characteristics at each time point were performed using one-way ANOVA or Kruskal-Wallis tests for continuous/ordinal variables, and Pearson’s chi-square tests for categorical variables.

Cumulative incidence and incidence rates of clinical outcomes were estimated for predefined intervals. To explore time-dependent risk patterns, landmark analyses were performed at 3 months, 1 year, 2 years, and 3 years. For each interval, only patients who were event-free at the preceding time point were included. The first 3-month interval was further subdivided at 2 weeks to capture early trends. Cumulative incidence was estimated using the Kaplan–Meier method, and incidence rates were expressed as events per 100 person-months with 95% confidence intervals. Kaplan–Meier curves and log-rank tests were used to compare cumulative event rates across discharge treatment groups.

To enhance interpretability, treatment groups were consolidated into 5 categories for outcome comparisons: (1) NOAC monotherapy, (2) VKA or other anticoagulants, (3) antiplatelet-only (combining single and dual agents, with separate analysis for each within the first 48 hours), (4) combination therapy, and (5) no treatment.

Subgroup analyses for the primary outcome were prespecified by age, sex, presence of symptomatic steno-occlusion, and coronary heart disease.

To evaluate associations between treatment and clinical events, multivariable Cox proportional hazards models were constructed separately for initial therapy (within 48 hours, related to in-hospital events) and discharge therapy (related to post-discharge events). Incidence rates for in-hospital events were calculated per 1,000 person-hours. All models adjusted for age, sex, NIHSS score, premorbid mRS, hypertension, diabetes, coronary heart disease, prior stroke or TIA, and symptomatic steno-occlusion. NOAC monotherapy served as the reference group.

All statistical tests were two-sided, with P values <0.05 considered statistically significant. Analyses were conducted using SAS version 9.4 (SAS Institute, Cary, NC) and R version 4.2.1 (R Foundation for Statistical Computing, Vienna, Austria).

### Ethics and Approvals

The study protocol was approved by the Institutional Review Boards of all participating centers. Written informed consent was obtained from all participants or their legal representatives. The study was conducted in accordance with the Declaration of Helsinki.

### Data Availability

The data that support the findings of this study are available from the corresponding author upon reasonable request. Deidentified participant data may be shared with qualified researchers for non-commercial academic purposes, subject to institutional and ethical approvals.

## Results

Among 2,965 patients with AF-related AIS, the mean (SD) age was 75.3 (10.2) years, and 54.1% were men. The median follow-up duration was 1.92 years (IQR 0.99-2.08). Valvular AF was present in 1.8% of participants. At baseline, the median NIHSS score was 6 (IQR 2-14), and 70.2% had hypertension (Table S1). Symptomatic steno-occlusion of relevant arteries was observed in 55.6%, coronary heart disease in 15.2%, and had a prior history of stroke or TIA in 25.4%. Prior to the index stroke, 30.1% were receiving antiplatelet therapy, 20.0% NOACs, and 6.8% VKAs.

Marked heterogeneity in antithrombotic strategies was observed during the acute phase. Within 48 hours of admission, 29.4% received single antiplatelet therapy, 21.5% dual antiplatelet therapy, and 23.2% NOAC monotherapy (Figure 1 and Table S2). At discharge, a substantial shift toward NOAC monotherapy (65.7%) was observed, and this pattern remained stable during follow-up, with 76.8% of patients (excluding those with missing data) still on NOAC therapy at 3 years. The proportion of patients on combined therapy with antiplatelet and anticoagulant agents peaked at discharge (13.7%) and declined to 6.4% by the second year.

**Figure 1.**
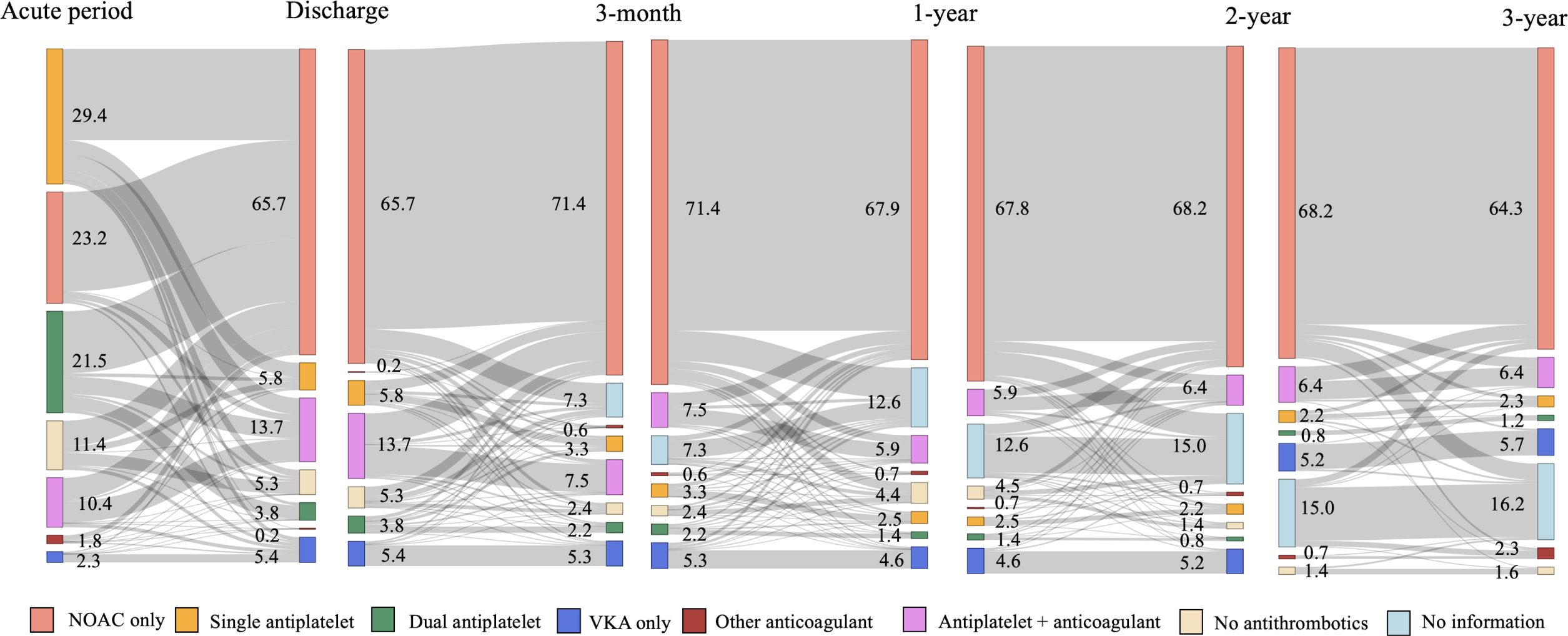
Temporal Changes in Antithrombotic Strategies From Acute Period to 3-Year Follow-up. Sankey diagram showing transitions in antithrombotic therapy across predefined time points. The width of the flow bands represents the proportion of patients on each therapy. NOAC indicates non-vitamin K antagonist oral anticoagulant; VKA, vitamin K antagonist. The acute period was defined as within 48 hours after admission. Antithrombotic strategies were categorized as NOAC only, VKA only, other anticoagulant, single antiplatelet, dual antiplatelet, antiplatelet + anticoagulant, no antithrombotics, and no information.

Among 2252 patients prescribed NOAC therapy at discharge, 29.8% had received single antiplatelet therapy, 20.2% dual antiplatelet therapy, and 32.5% NOACs within the first 48 hours (Figure 1 and Table S2). A notable proportion of patients initially treated with antiplatelet agents were administered combination therapy with antiplatelets and anticoagulants at discharge—11% of single and 17% of dual antiplatelet users. About 40% of those initially on combination therapy remained on it at discharge, but most transitioned to NOAC monotherapy during follow-up. VKA use remained stable at approximately 5.4%. Apixaban was the most prescribed NOAC therapy, followed by edoxaban (Table S3). Among 2,446 patients completing 1-year follow-up, 2,172 (88.8%) provided adherence data. These patients reported high medication compliance, taking their prescribed antithrombotic therapy for a mean of 97.5% of days in the preceding month.

Patients discharged on single antiplatelet agents or no antithrombotic therapy had more severe strokes (median NIHSS, 12), and higher rates of symptomatic steno-occlusion (68.3% and 73.2%, respectively) (Table). Patients discharged on single antiplatelet agents or no antithrombotic therapy had more severe strokes (median NIHSS, 12) and higher rates of symptomatic steno-occlusion (68.3% and 73.2%, respectively) (Table). This severity-based treatment selection was evident from the acute phase and persisted during follow-up, with differences less pronounced at 2 and 3 years (Tables S4-S9). The observed pattern likely reflects clinical concerns about hemorrhagic transformation risk in patients with large infarcts, leading to conservative anticoagulation strategies despite the known benefits of stroke prevention in AF patients. In contrast, those discharged on combination therapy had milder strokes (median NIHSS, 3), but a higher prevalence of diabetes (41.3%), coronary heart disease (31.1%), peripheral artery disease (3.6%), and prior history of stroke or TIA (36.2%). Notably, 41.3% had received anticoagulant therapy prior to the index stroke..

**Table.** Baseline characteristics according to antithrombotic strategy at discharge.

|  | NOAC only<br>(N = 1,882) | VKA only<br>(N = 155) | Other<br>anticoagulant<br>(N = 6) | Single<br>Antiplatelet<br>(N = 167) | Dual antiplatelet<br>(N = 108) | Antiplatelet +<br>anticoagulant<br>(N= 392) | No<br>antithrombotics<br>(N = 153) | P-value<sup>a)</sup> |
| --- | --- | --- | --- | --- | --- | --- | --- | --- |
| Age, mean (SD) | 75.3 (10.1) | 69.6 (11.4) | 68.0 (12.5) | 76.5 (10.4) | 76.4 (10.5) | 75.6 (8.7) | 75.2 (10.5) | <0.001 |
| Male sex, N (%) | 1049 (55.7) | 50 (32.3) | 3 (50.0) | 84 (50.3) | 59 (54.6) | 236 (60.2) | 77 (50.3) | <0.001 |
| Onset to arrival time, min, median (IQR) | 175.5 (59, 720) | 130 (52, 690) | 238 (25, 477) | 171 (69, 702) | 273.5 (90, 1079) | 311 (85.5, 129) | 139 (60, 462) | <0.001 |
| Premorbid mRS, median (IQR) | 0 (0, 1) | 0 (0, 1) | 0 (0, 4) | 0 (0, 1) | 0 (0, 1) | 0 (0, 1) | 0 (0, 1) | 0.648 |
| Initial NIHSS, median (IQR) | 6 (2, 14) | 6 (2, 16) | 6 (4, 13) | 12 (5, 18) | 5 (2, 13.5) | 3 (1, 9) | 12 (5, 17) | <0.001 |
| Risk factors, N (%) |  |  |  |  |  |  |  |  |
| Hypertension | 1314 (69.8) | 94 (60.6) | 2 (33.3) | 110 (65.9) | 92 (85.2) | 291 (74.2) | 108 (70.6) | <0.001 |
| Diabetes | 537 (28.5) | 41 (26.5) | 2 (33.3) | 53 (31.7) | 39 (36.1) | 162 (41.3) | 57 (37.3) | <0.001 |
| Hyperlipidemia | 509 (27.0) | 32 (20.6) | 2 (33.3) | 44 (26.3) | 35 (32.4) | 128 (32.7) | 35 (22.9) | 0.057 |
| Coronary heart disease | 227 (12.1) | 20 (12.9) | 0 (0.0) | 27 (16.2) | 22 (20.4) | 122 (31.1) | 21 (13.7) | <0.001 |
| Peripheral artery disease | 28 (1.5) | 2 (1.3) | 1 (16.7) | 2 (1.2) | 4 (3.7) | 14 (3.6) | 4 (2.6) | <0.001 |
| Previous stroke or TIA | 443 (23.5) | 43 (27.7) | 2 (33.3) | 35 (21.0) | 24 (22.2) | 142 (36.2) | 39 (25.5) | <0.001 |
| Current smoking | 241 (12.8) | 12 (7.7) | 1 (16.7) | 18 (10.8) | 12 (11.1) | 61 (15.6) | 18 (11.8) | 0.288 |
| Reperfusion therapy, N (%) |  |  |  |  |  |  |  |  |
| IV thrombolysis | 352 (18.7) | 19 (12.3) | 1 (16.7) | 26 (15.6) | 8 (7.4) | 38 (9.7) | 45 (29.4) | <0.001 |
| Endovascular thrombectomy | 456 (24.2) | 49 (31.6) | 2 (33.3) | 43 (25.7) | 18 (16.7) | 55 (14.0) | 48 (31.4) | <0.001 |
| Prior medication, N (%) |  |  |  |  |  |  |  |  |
| Antiplatelet | 511 (27.2) | 34 (21.9) | 1 (16.7) | 41 (24.6) | 59 (54.6) | 171 (43.6) | 48 (31.4) | <0.001 |
| VKA | 69 (3.7) | 75 (48.4) | 3 (50.0) | 6 (3.6) | 3 (2.8) | 30 (7.7) | 6 (3.9) | <0.001 |
| NOAC | 378 (20.1) | 13 (8.4) | 2 (33.3) | 26 (15.6) | 6 (5.6) | 132 (33.7) | 24 (15.7) | <0.001 |
| Statin | 494 (26.2) | 44 (28.4) | 1 (16.7) | 38 (22.8) | 34 (31.5) | 143 (36.5) | 34 (22.2) | <0.001 |
| Discharge statin, N (%) | 1685 (89.5) | 122 (78.7) | 3 (50.0) | 139 (83.2) | 100 (92.6) | 369 (94.1) | 74 (48.4) | <0.001 |
| Symptomatic steno-occlusion of the relevant<br>arteries, N (%) | 993 (52.8) | 78 (50.3) | 4 (66.7) | 114 (68.3) | 56 (51.9) | 216 (55.1) | 112 (73.2) | <0.001 |
| Valvular atrial fibrillation, N (%) | 6 (0.3) | 37 (23.9) | 0 (0.0) | 0 (0.0) | 2 (1.9) | 4 (1.0) | 4 (2.6) | <0.001 |

a) p-values by one-way ANOVA, Pearson's chi-square test, and Kruskal-Wallis test as appropriate
IQR, interquartile range; mRS, modified Rankin Scale; NIHSS, National Institutes of Health Stroke Scale; NOAC, non-vitamin K antagonist oral anticoagulant; SD, standard deviation; TIA, transient ischemic attack; VKA, vitamin K antagonist.

During the median follow-up of 1.92 years, the 3-year cumulative incidence of the primary outcome was 40.7%. For bleeding events, the 3-year cumulative incidence was 10.7% for any bleeding and 4.8% for major bleeding (BARC type 3 or 5) (Figure 2 and Table S10). The primary outcome demonstrated a distinct temporal pattern, with the highest incidence rate during the first 2 weeks (32.70 events per 100 person-months), declining to 7.92 events per 100 person-months over the first 3 months, and further decreasing thereafter (0.81 between 3 months and 1 year) (Figure S1 and Table S11). Similar trends were observed for stroke recurrence, all-cause mortality, and any bleeding (Figure 2 and Table S10).

**Figure 2.**
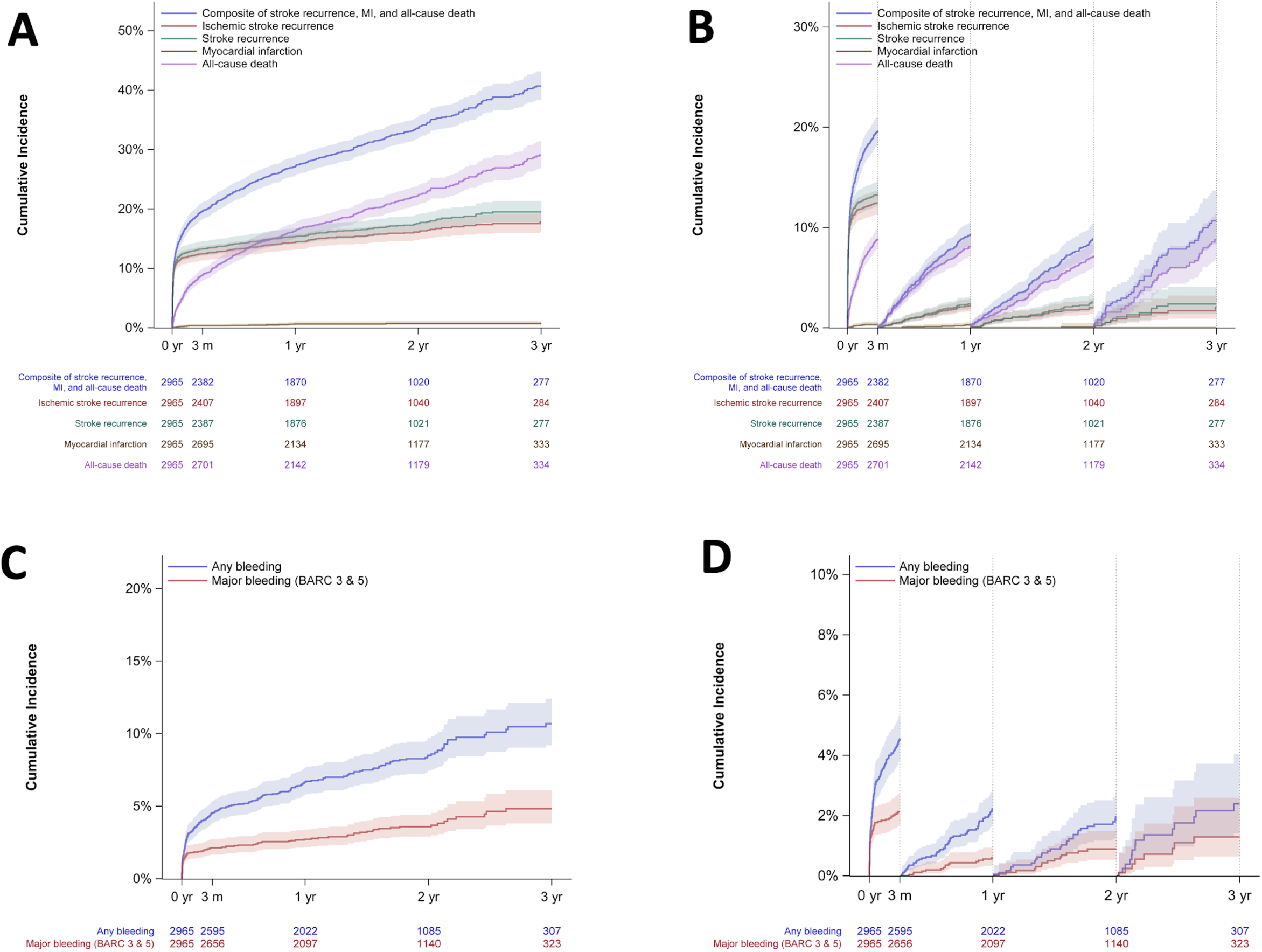
Cumulative Incidence of Primary and Secondary Outcomes Over 3-Year Follow-up. A, Overall cumulative incidence curves for the primary composite outcome (recurrent stroke, myocardial infarction, and all-cause death), and individual components including ischemic stroke recurrence. B, Landmark analysis for the primary composite outcome (recurrent stroke, myocardial infarction, and all-cause death), and individual components including ischemic stroke recurrence at 3-month, 1-year, 2-year, and 3-year time points showing conditional cumulative incidence rates. C, Overall cumulative incidence curves for bleeding events. D, Landmark analysis for bleeding events at 3-month, 1-year, 2-year, and 3-year time points showing conditional cumulative incidence rates. Shaded areas represent 95% confidence intervals. Numbers below graphs indicate patients at risk at each time point.

Subgroup analyses revealed that older age (75 years or above), female sex, coronary heart disease, and symptomatic steno-occlusion were associated with higher cumulative incidence of the primary outcome (log-rank P<0.05 for all comparisons) (Figure S2).

Based on discharge antithrombotic treatment strategy, the 3-month incidence rate of the primary outcome was highest among patients without antithrombotic therapy (18.44 [95% CI 14.26-23.86] per 100 person-months), followed by antiplatelet-only therapy (11.98 [95% CI 9.57-15.01]), and lowest in those receiving NOAC monotherapy (4.95 [95% CI 4.37-5.61]). These differences persisted throughout the follow-up period: the 3-year cumulative incidence was 63.0% for no antithrombotic therapy, 47.9% for antiplatelet-only therapy, and 35.7% for NOAC therapy (Figure 3A, B and Table S12). Secondary outcomes showed similar trends (Figures S3, S4 and Tables S13 to S18).

**Figure 3.**
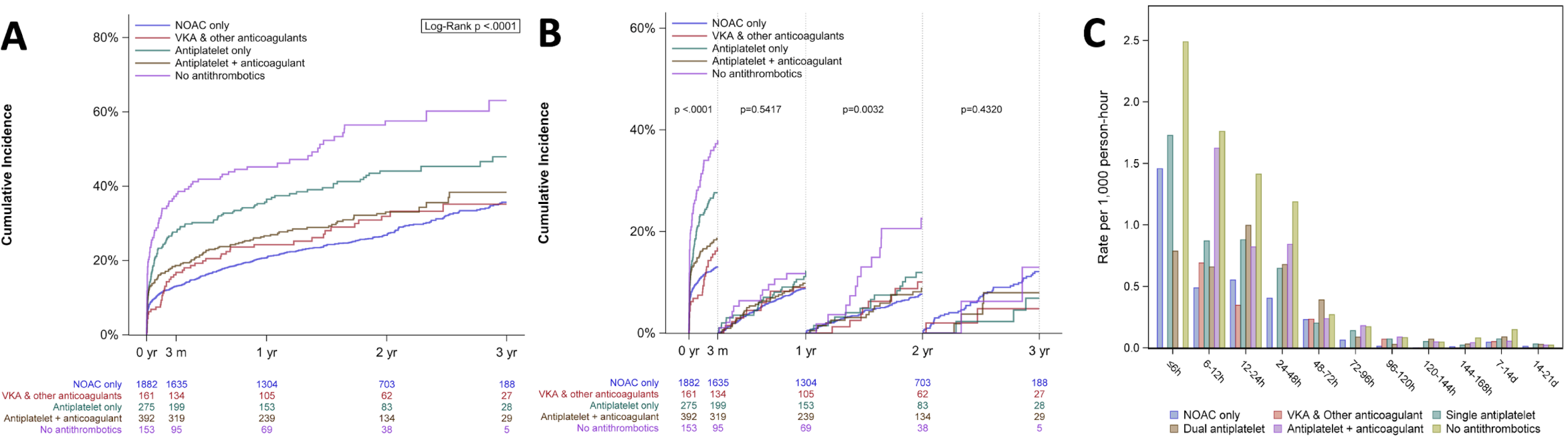
Event Rates According to Different Antithrombotic Strategies. A, Cumulative incidence curves for the primary outcome stratified by discharge antithrombotic strategy. B, Landmark analysis showing conditional cumulative incidence rates by discharge antithrombotic strategy. C, Incidence rates (per 1,000 person-hours) during hospitalization according to initial antithrombotic strategy. P values are from log-rank tests. Numbers below graphs indicate patients at risk at each time point.

Regarding antithrombotic treatment within first 48 hours, in-hospital event rates (per 1,000 person-hours) were highest among patients who received no therapy, followed by those on combination or antiplatelet-only regimens, and lowest in those receiving NOAC monotherapy (Figure 3C and Figure S5).

In multivariable analyses adjusting for relevant covariates, initial treatment with dual antiplatelet therapy (hazard ratio [HR] 1.65, 95% confidence interval [CI] 1.08-2.30), combination therapy (1.75 [1.18-2.60]), and no therapy (1.65 [1.05-2.19]) was associated with significantly higher in-hospital risk of the primary outcome compared with NOAC monotherapy. At discharge, when compared with NOAC monotherapy, no antithrombotic therapy was associated with a 2-fold increased risk of the primary outcome during follow-up (HR 2.05, 95% CI 1.60-2.62), while antiplatelet-only therapy (HR, 1.60 [95% CI, 1.31-1.96]) and VKA or other oral anticoagulants (HR, 1.36 [95% CI, 1.01-1.83]) were also associated with elevated risks (Figure 4).

**Figure 4.**
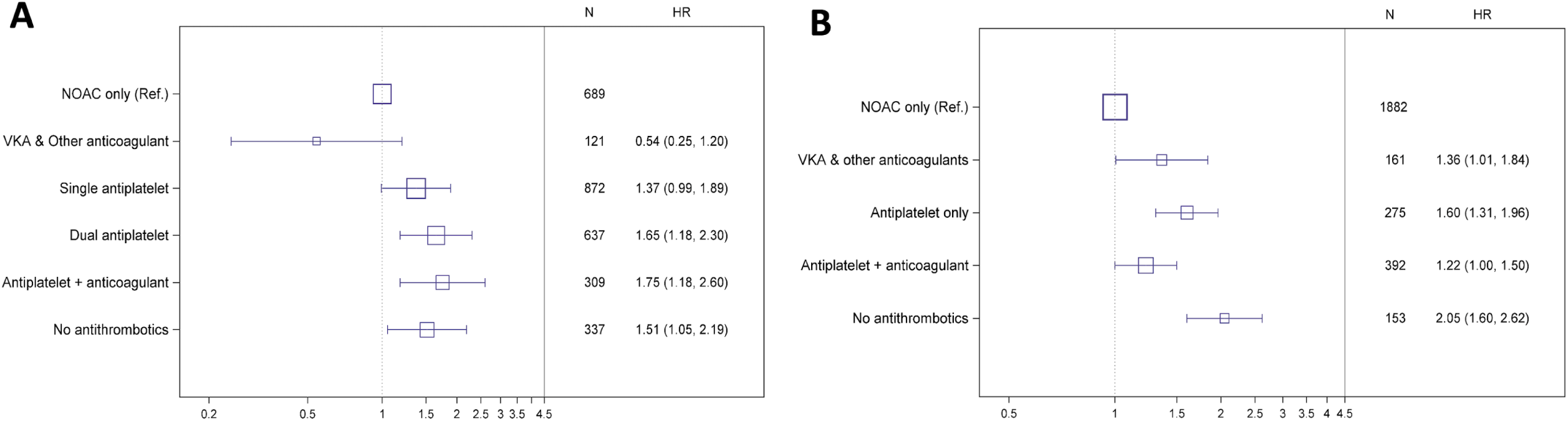
Adjusted Hazard Ratios for Primary Outcome by Antithrombotic Strategy. Forest plots showing adjusted hazard ratios with 95% confidence intervals for the primary outcome according to antithrombotic strategy: A, initial therapy within the first 48 hours and its association with in-hospital outcomes; B, discharge therapy and its association with post-discharge outcomes. NOAC monotherapy served as the reference group. Models were adjusted for age, sex, initial NIHSS score, premorbid mRS, hypertension, diabetes, coronary heart disease, previous stroke/TIA, and symptomatic steno-occlusion. NIHSS indicates National Institutes of Health Stroke Scale; mRS, modified Rankin Scale; TIA, transient ischemic attack.

## Discussion

In this large prospective cohort of patients with AF-related AIS, we identified several key findings regarding contemporary antithrombotic practices and associated outcomes. First, substantial heterogeneity was observed in acute phase antithrombotic strategies, with nearly half of patients receiving antiplatelet therapy alone and only 23.2% receiving NOAC monotherapy. This variability diminished by discharge, with NOAC monotherapy becoming predominant (65.7%) and remaining stable over time. Second, adverse clinical events were highly concentrated in the early post-stroke period, particularly within the first two weeks—coinciding with the phase of greatest treatment heterogeneity. Third, antithrombotic regimens at both the acute and discharge stages were associated with clinical outcomes, with NOAC therapy consistently associated with lower event rates compared with antiplatelet-only therapy or no treatment.

This early treatment heterogeneity occurring during a period of heightened vulnerability to recurrent events, suggests a critical gap between clinical need and the implementation of evidence-based care. Patients with more severe strokes were less likely to receive anticoagulant therapy and experienced higher rates of adverse outcomes, underscoring the challenges of optimal management in these high-risk individuals.

Compared to recent clinical trials, our cohort exhibited a higher rate of early events. For instance, the ELAN trial reported a 30-day cumulative incidence of 3.5% following randomization,^9^ whereas the cumulative incidence in our cohort reached 5.6% within the first two weeks after stroke onset. This discrepancy likely reflects the real-world nature of our study population and its Asian ancestry. Specifically, our study included a higher proportion of patients with symptomatic steno-occlusion of relevant cerebral arteries (55.6%)—a characteristic more commonly observed in Asian populations (Figure S3).^21,22^

The high frequency of early events poses a significant therapeutic challenge. Although current guidelines often recommend delaying anticoagulation initiation, particularly in patients with large infarcts, due to concerns about hemorrhagic transformation, recent trials support early NOAC initiation.^1,23,24^ The TIMING trial demonstrated that NOAC initiation within 4 days was noninferior to delayed initiation (5-10 days), while the ELAN trial showed that severity-guided early initiation reduced vascular events without increasing hemorrhagic risk.^9,10^ Our findings reinforce the importance of re-evaluating the optimal timing of anticoagulation initiation.

The severity-based treatment selection observed in our study, where patients with more severe strokes were less likely to receive anticoagulation, highlights the need for safer therapeutic alternatives. Emerging agents such as factor XIa inhibitors may offer a safer alternative for patients at high risk of bleeding.^25^ The PACIFIC-AF trial showed that asundexian was associated with a lower bleeding risk than apixaban, while maintaining comparable antithrombotic efficacy.^26^ These agents may prove valuable for patients deemed ineligible for currently used anticoagulant therapies.

Several earlier cohort studies have explored antithrombotic use in AF-related stroke. The SAMURAI-NVAF and K-ATTENTION studies, conducted between 2011 and 2015, reported lower rates of NOAC therapy administration at discharge (41.7% and 26.9%, respectively) compared with our cohort (65.7%).^27,28^ The Berlin Atrial Fibrillation Registry (2014-2016) showed similar adoption rates (68.1%) to our study, reflecting the global trend towards use of NOACs.^29^ While earlier studies suggested comparable outcomes between NOAC and VKA therapy users, our findings demonstrate more favorable outcomes with NOACs, likely reflecting improvements in both patient selection for NOACs and stroke care practices in contemporary settings.^30^ Notably, patients discharged on VKAs experienced higher rates of adverse events, despite multivariable adjustment—likely due to well-documented limitations of VKA therapy, including difficulty in maintaining adequacy of time in therapeutic range and the need for frequent INR monitoring.^2,31^ These findings highlight the need for safer and more practical alternatives in patients who are ineligible for NOAC therapies but remain on VKAs.

A substantial proportion of patients received combination therapy with antiplatelets and anticoagulants, particularly those with coronary artery disease or symptomatic cerebral arterial steno-occlusion. Although current guidelines recommend short-term combination therapy in the setting of acute coronary syndrome,^32,33^ optimal strategies for patients with AF and concomitant atherosclerotic disease after stroke need to be clarified. The elevated event rates in patients with concomitant coronary artery disease or symptomatic steno-occlusion of relevant cerebral arteries (Figure S2C and S2D) suggest that the current approach may be inadequate for these subgroups of patients.

This study has several limitations. First, because the cohort originated from tertiary centers in Korea and the participants were of Asian ancestry, the findings may not be generalizable to other populations or healthcare systems. However, consistency with national stroke audit data supports the broader applicability of our findings, at least in Korea.^34^ Second, approximately 13% of eligible patients did not provide consent and were excluded from the study, which may introduce selection bias. Third, as an observational study, causal inferences regarding treatment effectiveness are limited, and residual confounding cannot be excluded. Fourth, while medication adherence assessed at 1-year follow-up showed high compliance rates (97.5%), this relied on self-reported data from a single time point with 11.2% missing responses, potentially overestimating actual adherence.

In conclusion, this large, real-world cohort of patients from Korea with AF-related AIS demonstrates that early post-stroke adverse events were frequent and associated with initial and discharge choice of antithrombotic strategies. The concentration of risk in the early phase after AIS underscores the need for future trials targeting this critical window. Additionally, the suboptimal outcomes in patients not receiving anticoagulation highlight the need for safer, more personalized treatment options. Efforts to optimized early antithrombotic management, especially with NOAC therapies may improve outcomes in this high-risk population.

## Nonstandard Abbreviations and Acronyms

AF: Atrial fibrillation
AIS: Acute ischemic stroke
NOAC: Non–vitamin K antagonist oral anticoagulant
VKA: Vitamin K antagonist
CRCS-K: Clinical Research Collaboration for Stroke in Korea
NIHSS: National Institutes of Health Stroke Scale
mRS: Modified Rankin Scale
TIA: Transient ischemic attack
BARC: Bleeding Academic Research Consortium

